# COVID-19 wastewater surveillance in rural communities: Comparison of lagoon and pumping station samples

**DOI:** 10.1101/2021.05.01.21256458

**Authors:** Patrick M. D’Aoust, Syeda Tasneem Towhid, Élisabeth Mercier, Nada Hegazy, Xin Tian, Kamya Bhatnagar, Zhihao Zhang, Colleen C. Naughton, Alex E. MacKenzie, Tyson E. Graber, Robert Delatolla

**Affiliations:** Department of Civil Engineering, University of Ottawa, Ottawa, Ontario, Canada; Children’s Hospital of Eastern Ontario Research Institute, Ottawa, Ontario, Canada; University of California Merced, Merced, California, United States

**Keywords:** wastewater treatment lagoon, SARS-CoV-2, wastewater-based epidemiology, wastewater surveillance, COVID-19, pepper mild mottle virus

## Abstract

Wastewater-based epidemiology/wastewater surveillance has been a topic of significant interest over the last year due to its application in SARS-CoV-2 surveillance to track prevalence of COVID-19 in communities. Although SARS-CoV-2 surveillance has been applied in more than 50 countries to date, the application of this surveillance has been largely focused on relatively affluent urban and peri-urban communities. As such, there is a knowledge gap regarding the implementation of reliable wastewater surveillance in small and rural communities for the purpose of tracking rates of incidence of COVID-19 and other pathogens or biomarkers. This study examines the relationships existing between SARS-CoV-2 viral signal from wastewater samples harvested from an upstream pumping station and from an access port at a downstream wastewater treatment lagoon with the community’s COVID-19 rate of incidence (measured as percent test positivity) in a small, rural community in Canada. Real-time quantitative polymerase chain reaction (RT-qPCR) targeting the N1 and N2 genes of SARS-CoV-2 demonstrate that all 24-hr composite samples harvested from the pumping station over a period of 5.5 weeks had strong viral signal, while all samples 24-hr composite samples harvested from the lagoon over the same period were below the limit of quantification. RNA concentrations and integrity of samples harvested from the lagoon were both lower and more variable than from samples from the upstream pumping station collected on the same date, indicating a higher overall stability of SARS-CoV-2 RNA upstream of the lagoon. Additionally, measurements of PMMoV signal in wastewater allowed to normalize SARS-CoV-2 viral signal for fecal matter content, permitting the detection of actual changes in community prevalence with a high level of granularity. As a result, in sewered small and rural communities or low-income regions operating wastewater lagoons, samples for wastewater surveillance should be harvested from pumping stations or the sewershed as opposed to lagoons.

## 1. Introduction

In late 2019, cases of COVID-19 began to spread rapidly internationally (Li et al., 2020). It became clear to public health officials across the world that this new disease, caused by the severe acute respiratory syndrome coronavirus 2 (SARS-COV-2) (Eurosurveillance Editorial Team, 2020), was rapidly becoming a pandemic-potential pathogen due to its relatively low virulence but high degree of infectiousness (He et al., 2020). More than a year after the first cases of COVID-19, the world is still grappling with the disease and newer and more infectious variants of the virus are spreading (Duong, 2021; Galloway et al., 2021; Volz et al., 2020; Walensky et al., 2021), causing widespread disease and death (WHO COVID-19 Dashboard https://covid19.who.int/). At the time of writing (May 20^th^, 2021), more than 2.4% of the global population (191.1 M) has been infected by SARS-CoV-2, and 2.1% of those infected have died (4.1 M) (WHO COVID-19 Dashboard).

Now widely applied in over 1,000 sites in more than 50 countries worldwide (Ahmed et al., 2020; Arora et al., 2020; Bivins et al., 2020b; D’Aoust et al., 2021a; Gonzalez et al., 2020; Mao et al., 2020; Medema et al., 2020; Naughton et al., 2021; Polo et al., 2020; Randazzo et al., 2020; Sims and Kasprzyk-Hordern, 2020; Thompson et al., 2020), Wastewater surveillance (WWS) efforts conducted with RT-qPCR are underway around the world, focused primarily in larger metropolitan areas of higher income countries (Bivins et al., 2020b). By and large rural communities and low-income countries have not had the same services afforded to them as urban and peri-urban communities (WEF Network of Wastewater-Based Epidemiology, 2021) and higher income countries, which is based on numerous factors that include but are not limited to: i) discrepancies in financial and material resources, ii) distance to research, academic and governmental facilities capable of carrying out the analyses and iii) capacity of the local public health unit to take in the results and act upon them. Furthermore, smaller, rural communities and low-income countries without larger mechanical water resource recovery facilities may not have the staff, equipment or expertise to carry out sampling for SARS-CoV-2 viral detection in wastewater (Haider et al., 2016; Naughton et al., 2021; Switzer et al., 2016). Larger facilities are often equipped with automatic composite samplers throughout the plant, making the implementation of a WWS monitoring program in comparison relatively easy for routine purposes. Furthermore, there is a growing consensus that higher concentrations of SARS-CoV-2 viral particles are found in wastewater solids (Chik et al., 2021; D’Aoust et al., 2021b; Graham et al., 2020; Peccia et al., 2020; Pecson et al., 2021) and as such several WWS efforts are now focusing on measuring signal from solid fractions of samples (Chik et al., 2021; Wolfe et al., 2021). In small and rural communities, harvesting solids may be problematic as dedicated solid separation units present in larger facilities located in urban and peri-urban communities may not exist in smaller facilities, requiring different sampling approaches. Unfortunately, the limited resources available in small and rural communities or lower income countries often compromise the WWS efforts. The communities may in turn be at the mercy of funding or mandates due to the lack physical, material and/or financial resources. As an example of the precariousness of WWS in smaller communities, Finnish authorities recently announced in June 2021 that they would-be discontinuing SARS-CoV-2 WWS efforts in cities with populations smaller than 150,000 (Finnish institute for health and welfare, 2021). In contrast, remote communities in the Northwest Territories of Canada have implemented wastewater surveillance to monitor the communities for COVID-19 viral signal (Government of Northwest Territories, 2021), despite the low population of the geographic area (<45,000). Having clear guidelines, appropriate analytical methods and low-cost strategies for surveillance which are also applicable for smaller communities will therefore be critical to ensure that WWS efforts service the most residents in each region.

Most WWS efforts attempt to predict trends of epidemiological metrics of COVID-19 in the general population by quantifying increases and decreases in the rates of clinical cases of COVID-19 (Bivins et al., 2020a; D’Aoust et al., 2021a; Hill et al., 2020; Kumar et al., 2021; Polo et al., 2020). These environmental studies will often focus on collecting samples within the raw influent or primary sludge due to the relatively high concentration of solids in these wastewater streams (Hill et al., 2020). However, several municipalities operating smaller types of treatment systems such as waste stabilization ponds, also known as wastewater treatment lagoons, do not have direct access to raw influent or primary sludge. Smaller communities may however have direct access to the waste stabilization ponds and pumping/lift stations (MOE Ontario, 2008). Wastewater treatment lagoons are commonly used in the world, with over 1,200 in operation in Canada (Statistics Canada, 2016), over 5,500 in Europe (Mara, 2009), and over 8,000 in the United States alone (USEPA, 2011). Solids separation occurs in lagoons due to the slowing of flow velocities, leading to particle settling, particularly in the same area of the lagoon that oxidizes carbonaceous deleterious substances or in lagoon areas or isolated lagoon units designed specifically for solids sedimentation (Asano et al., 2007; D’Aoust et al., 2021c; Leblond et al., 2020). As such, harvesting of wastewater solids in lagoon treatment systems with the goal of performing WWS is very difficult due to potentially long retention times in lagoons, the degradation of RNA targets due to environmental temperature fluctuations and the difficulty of collecting “fresh” solids from a lagoon representing current incidence of COVID-19 in the community. Furthermore, as lagoon systems are located outdoors and exposed to ambient temperatures, in locales where air temperatures can dip below freezing these systems may become difficult to sample due to the presence of ice-cover. High temperatures during summer months and UV radiation from sunlight may also further degrade viral RNA (Verbyla et al., 2017). As a result, smaller communities may not have evident sampling locations to collect wastewater samples containing SARS-CoV-2 viral particles which can accurately represent changes in prevalence of the disease in the community. Another factor easing the implementation of WWS programs in small and rural communities is to see if the community is sewered or not. In communities that are not sewered and where centralized sampling points are not available, sewage brought to lagoons can be sampled from sewage and sludge trucks while they are being emptied at the facility (Telliard, 1989).

In preparation for applying SARS-CoV-2 wastewater surveillance initiatives in a small sewered community in Eastern Ontario (Canada), wastewater samples were collected from an access/sampling point situated between the first and second cells of a lagoon treatment system, and from the last pumping station on the sewer network located upstream of the lagoon treatment system. The specific objectives of this study were to: i) compare SARS-CoV-2 signal at both sampling locations for strength of the RNA viral signal and RNA integrity and ii) compare the higher integrity longitudinal SARS-CoV-2 signal to existing community epidemiological data to ascertain the ability of WWS to track and predict and correlate with trends in rates of incidence of COVID-19 in small and rural communities.

## 2. Experimental methods

### Rural community sampling locations

The wastewater of a rural community of less than 5,000 inhabitants in Eastern Ontario (Canada) were sampled in this study between October 2020 and May 2021 (Figure 1). The rural community is sewered, with the wastewater flowing into the main pumping station located 1.3 km upstream of a wastewater treatment lagoon. The lagoon system consists of 3 cells/ponds (total surface area of approximately 182,000 m^2^) operated in-series that flow from cell #1 to #2 to #3. The lagoon receives continuous inflow with an average daily flow rate of 2,110 m^3^/d. The lagoon system discharges to a nearby river twice annually, in Spring (Mar. 7^th^ to May 15^th^) and Fall (Oct. 1^st^ to Dec. 19^th^). The lagoon system does not include a wetland component. Cell #3 has bottom-mounted aerators that are engaged prior to and during discharge to strip hydrogen sulfide before the release of treated wastewater to the natural environment. To control phosphorus concentrations in the final effluent of the lagoon treatment facility, a polyaluminum sulphate solution is injected directly into the pressurized wastewater pipe following the pumping station (force main) immediately before being released to the first cell of the lagoon system. The yearly average treatment efficiency of cBOD_5_, total suspended solids, total phosphorus, total ammonia nitrogen, total Kjeldahl nitrogen and alkalinity is of 96.0%, 94.9%, 98.0%, 96.9%, 94.1% and 64.6% removal efficiency, respectively.

**Figure 1:**
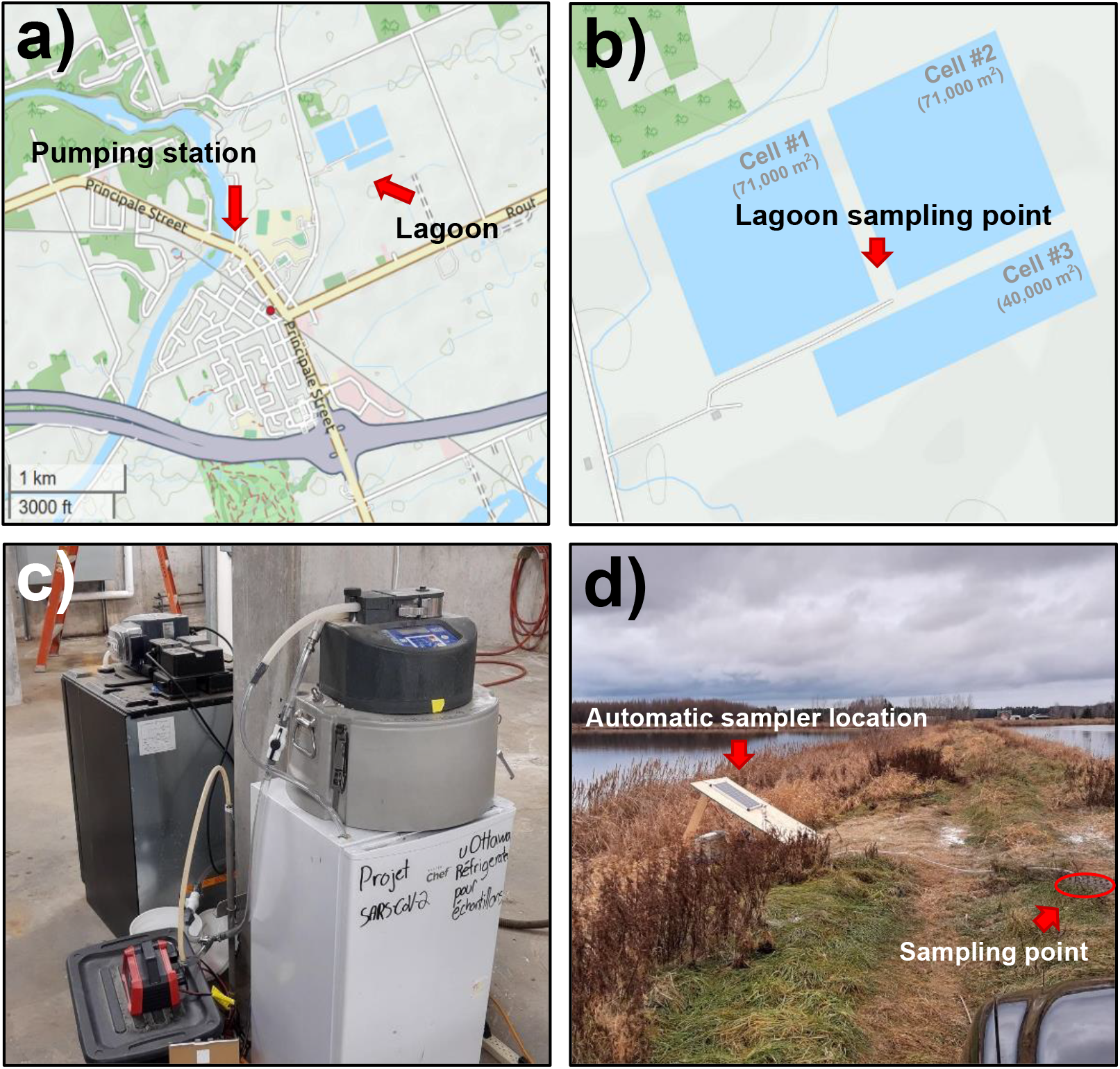
Sampling locations and configurations: a) geographic locations of pumping station and lagoon, b) lagoon treatment system consisting of three cells, identification of sampling point, c) automatic sampler at pumping station, sampler dispenses samples into refrigerator, and d) automatic sampler under the solar panel at lagoon sampling point and sampling point.

Two locations were sampled for wastewater in the rural community: i) the upstream pumping station, which receives the same annual volumetric flow of wastewater as the wastewater treatment lagoon system itself, and ii) an access/sampling point situated between cells 1 and 2 of the lagoon treatment system (Figure 1). The wastewater travel time between the pumping station and the inlet of the lagoon is approximately 15 minutes, while the residence time of the wastewater at the sampling location between the two cells of the lagoon ranges significantly due to the annual discharge design of the system (approximated residence times ranging between 80 hours and 10 days during the period of the study). The typical wastewater characteristics at the pumping station and at the lagoon effluent are shown in Table 1.

**Table 1.**
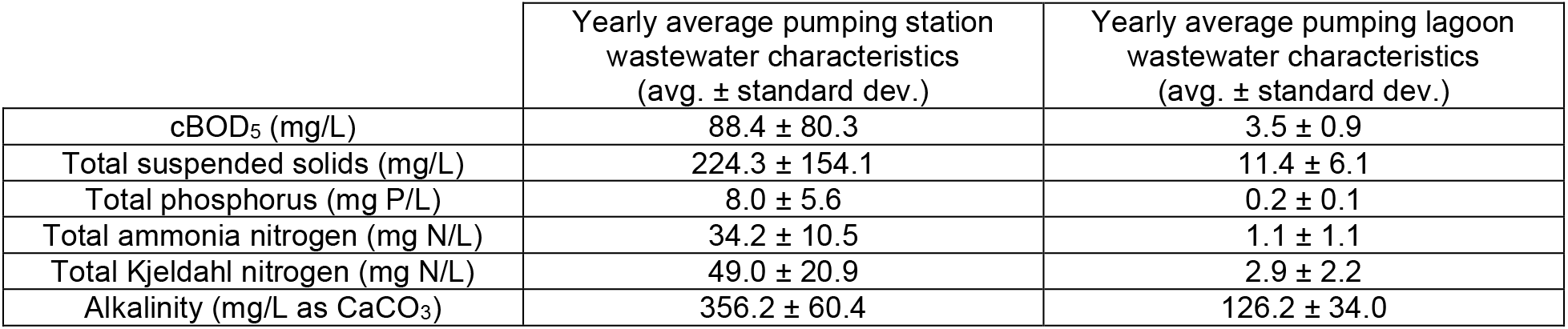
Yearly average wastewater characteristics at the pumping station and lagoon effluent.

### Sample collection

At the pumping station, 24-hr composite samples of wastewater were collected every 3 to 7 days from October 16^th^, 2020, to May 2^nd^, 2021, using an ISCO 6700 series automatic sampler (Teledyne ISCO, Lincoln, NE, USA). The autosampler located at the pumping station pumped the wastewater into a storage container located inside an adjacent refrigerator to maintain the samples at 4°C until collection (within 24 hours). At the lagoon sampling point, 24-hr composite samples of wastewater were also collected every 3 to 7 days from December 3^rd^, 2020, to January 11^th^, 2021. The autosampler located at the lagoon pumped the wastewater into a sealed bottle in an insulated foam container located adjacent to the autosampler. Outdoor temperatures at the lagoon during the study period (Dec. 3rd, 2020, to Jan. 11th, 2021) oscillated between -5.5°C and 1.3°C, hence allowing safe preservation of the samples between sampling and collection without additional refrigeration being required. composite samples were comprised of twenty-four 50 mL aliquots. During the pairwise comparison of both locations from Dec. 3rd, 2020, to Jan. 11th, 2021), the automatic samplers were programmed to collect samples at the same time at both locations. After every sampling cycle ended, the composite samples were collected and were transported on ice to the laboratory for analysis. Once in the laboratory, samples were concentrated immediately, and the resulting pellets were frozen at -30°C and processed within 14 days. After January 11^th^, site access became difficult due to low outdoor temperatures, freezing of the lagoon, and snowing conditions, making access to the automatic sampler highly difficult, which led to the cessation of sampling at the lagoon.

### Sample concentration, extraction, and PCR quantification

The composite samples were concentrated by allowing the samples to settle at 4°C for an hour, followed by the decantation of the supernatant to isolate the settled solids fraction. 40 mL of remaining solid fraction was then transferred to a 40 mL centrifuge tube and samples were then centrifuged for 45 mins at 10,000 x g at 4°C to isolate the centrifuged pellet. Sample pellets which could not be immediately processed were frozen at -30°C for a period of up to 14 days before being extracted. RNA was extracted and purified from the resulting pellet using the RNeasy PowerMicrobiome kit (Qiagen, Germantown, MD, USA) using a QIAcube Connect automated extraction platform, with the protocol modifications specified in an earlier study (D’Aoust et al., 2021b). The SARS-CoV-2 signal in the samples was assayed using a singleplex one-step RT-qPCR targeting the N1 and N2 gene regions of SARS-CoV-2 genome. The signal of pepper mild mottle virus (PMMoV) was also measured in each of the samples (samples were diluted 1/10 for measurements of PMMoV). In each PCR reaction, the reaction mix consisted of 1.5 µl of RNA template, 500 nM of each of forward and reverse primer (IDT, Kanata, Canada) in 4x TaqMan^®^ Fast Virus 1-step Mastermix (Thermo-Fisher, USA) with 125 nM probe (IDT, Kanata, Canada) in final volume of 10 µl. The samples were run in triplicates with non-template controls and were quantified using a five-point gradient of the EDX SARS-CoV-2 COV-19 RNA standard (Exact Diagnostics, USA). Reverse transcription (RT) was performed at 50°C for 5 minutes followed by RT inactivation and initial denaturation at 95°C for 20 seconds. This was followed by 45 cycles of denaturation at 95°C for 3 seconds and annealing/extension at 60°C for 30 seconds with a CFX Connect qPCR thermocycler (Bio-Rad, USA). The assay limit of detection (ALOD, ≥95% detection) was assessed (D’Aoust et al., 2021b) and determined to be approximately 2 copies/reaction for both N1 and N2 SARS-CoV-2 gene targets. The assay limit of quantification (ALOQ, CV=35%) was determined to be approximately 3.2 copies/reaction for N1 and 8.1 copies/reaction for N2 SARS-CoV-2 gene targets. SARS-CoV-2 N1 and N2 gene region viral signals were normalized by dividing the N1 and N2 gene copies per reaction by the PMMoV gene copies per reaction, as a means of normalizing the N1 and N2 signal by the quantity of fecal matter in the sample (D’Aoust et al., 2021b, 2021a; Graham et al., 2020; Kitajima et al., 2018). Vesicular stomatitis virus (VSV) was used as an internal control to monitor the efficiency of the viral concentration and extraction processes and was spiked into the sample prior to extraction. The extraction efficiency quantified with the VSV spike-in was between 3-4.5%. All samples were checked for inhibition by diluting the samples by a factor of 4 and 10 and measuring the corresponding drop in signal of PMMoV.

### Assessment of RNA Integrity

Samples were analysed for RNA integrity using an Agilent 2100 Bioanalyzer. RNA (2 µL) of each sample was loaded on an RNA 6000 Pico Chip (#5067-1513). Data analysis and RNA concentration calculations were performed using Agilent’s proprietary 2100 Expert software (version B.02.10.SI764).

### Collection of epidemiological data and correlation to wastewater viral signal

Weekly epidemiological data was obtained from the ICES COVID-19 dashboard (https://www.ices.on.ca/DAS/AHRQ/COVID-19-Dashboard) and the Eastern Ontario Health Unit (EOHU) dashboard (https://eohu.ca/en/covid/covid-19-status-update-for-eohu-region). Correlation analyses were then performed between the PMMoV-normalized SARS-CoV-2 viral signal in wastewater and the available epidemiological data.

## 3. Results & discussion

During the pairwise comparison of samples collected from both locations at the same time-point, all samples (5/5) from the pumping station showed detectable signal for the N1 and N2 gene regions of SARS-CoV-2 (Figure 2), as well as for PMMoV. Measurements of viral signal for the N1 gene region of the samples collected from the pumping station ranged from 5.5 × 10^3^ -4.6× 10^4^ genomic copies/L while measurements for the N2 gene region ranged from 4.3 × 10^3^ – 3.2 × 10^4^ genomic copies/L. Meanwhile, all composite samples collected at the lagoon sampling point between cells #1 and #2 of the lagoon were below the ALOQ and the ALOD for the N1 and N2 gene, and 4 out of 5 samples had no detectable SARS-CoV-2 viral signal altogether. During the same short span of side-by-side test period, PMMoV was only observed in three of the five lagoon samples.

**Figure 1:**
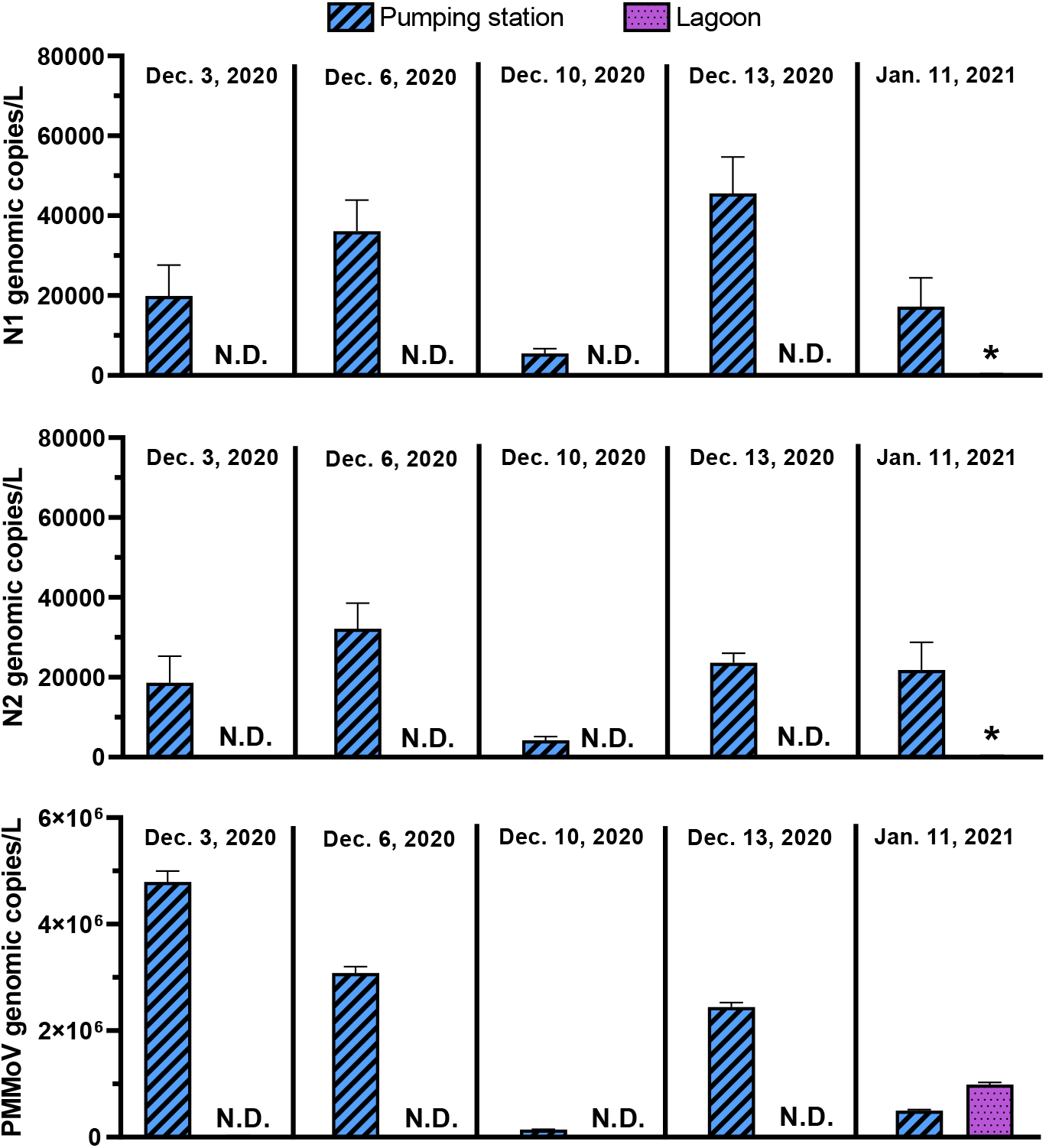
Comparison of genomic copies/L of pumping station samples and waste stabilization pond samples over time for the N1 and N2 SARS-CoV-2 gene regions, and PMMoV. Bars with a star (*) indicate that the sample signal was below the ALOQ.

In samples collected from the pumping station, measurements of PMMoV ranged from 1.4× 10^5^ – 4.8 × 10^6^ genomic copies/L, and in lagoon samples with PMMoV measurements, the PMMoV concentration ranged from 6.9 × 10^2^ – 9.9× 10^5^ genomic copies/L. The lack of PMMoV signal in some of the lagoon samples could potentially signify that near complete RNA degradation of PMMoV RNA occurred in the lagoon. When comparing viral signal measurements from the RT-qPCR analyses for the N1 and N2 SARS-CoV-2 gene regions and PMMoV between the pumping station and lagoon samples, pairwise comparisons clearly outline a stronger detection of both SARS-CoV-2 N gene regions and PMMoV viral signal in the samples collected from the pumping station (Figure 2). Furthermore, due to the ubiquity of PMMoV in wastewater, observing PMMoV measurements may allow to distinguish SARS-CoV-2 N1 and N2 gene region true non-detects from false-negatives caused by sample degradation or severe inhibition of the sample (Hong et al., 2021).

RNA of the SARS-CoV-2 and PMMoV targets in samples collected from the sampling location in the lagoon may have experienced more degradation due to extended residence times within the lagoon. Although temperature and storage time have been shown to affect SARS-CoV-2 viral signal degradation in wastewater (Hart and Halden, 2020), the similar storage temperature of the pumping station and lagoon samples during the pairwise study along with the short storage time until analysis enable this study to isolate the difference in signals to the conditions of the two sampling locations. Furthermore, due to the low temperature of the wastewater in the pumping station and within the lagoon during the study, viral degradation due to elevated temperatures is not a likely pathway of degradation. It is also possible that the RNA of the samples in the lagoon may have been subject to UV degradation once within the lagoon itself (Fongaro et al., 2012; Verbyla et al., 2017). Furthermore, as SARS-CoV-2 viral particles are believed to likely partition preferentially to wastewater solids in typical wastewater conditions (Arora et al., 2020; Chakraborty et al., 2021;

D’Aoust et al., 2021a; Graham et al., 2020; McLellan et al., 2021), a significant portion of the viral RNA may have settled throughout the first lagoon cell as the flow velocity of the wastewater immediately decreases upon entering the lagoon system’s first cell. Additionally, this facility doses polyaluminum sulphate for phosphorus abatement, which likely contributes to a more rapid and pronounced settling of solids. It is believed that the polyaluminum may also help flocculate SARS-CoV-2 genetic material, as evidenced by several studies employing aluminum-driven flocculation concentration methods (Barril et al., 2021; Randazzo et al., 2020). It is noted that all mechanisms contributing to potential degradation of viral signal in the lagoon are symptomatic of treatment of wastewater in the lagoon. Further investigations with a control location which does not use polyaluminum sulphate could be conducted to verify this hypothesis.

Total RNA concentrations were measured in a series of samples collected from the pumping station and the lagoon sampling point. Results of the analyses are shown in Figure 3. RNA concentrations are distinctly lower in lagoon samples as compared to pumping station samples. This is likely due to the presence of less fecally-associated biological material or greater degradation in the material collected in the samples collected in the lagoon.

**Figure 2:**
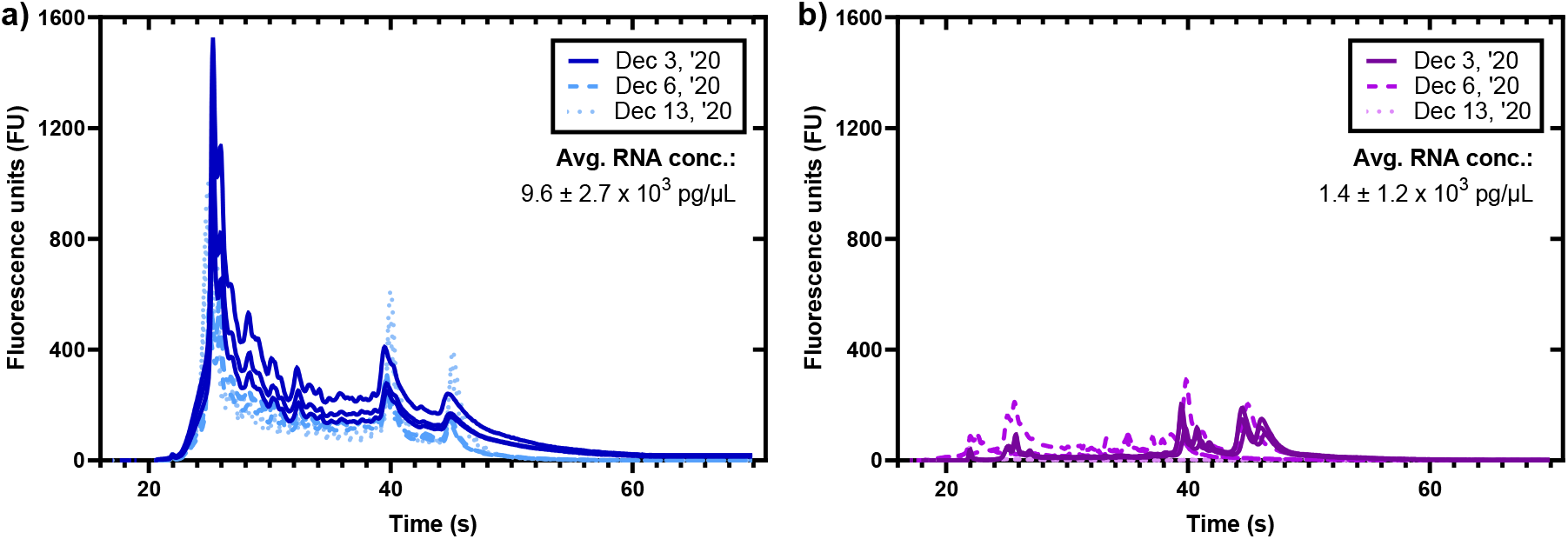
Electropherograms of samples from the a) pumping station and the b) lagoon, showing drastic differences in the RNA profiles.

The epidemiological data pertaining to this study were not available at the granulation of the studied community itself (pop.: ∼4,000; ∼5 km^2^), but rather for a larger geographical region (pop.: ∼200,000; ∼5,300 km^2^) that includes the community. The lack of available epidemiological data for the small community itself is indicative of the need for additional resources for small, rural communities and the potential for the application of WWS in these communities as an effective indicator of incidence of infections at the granulation of the community. Further, the clinical metric that was available at the geographic region level was percent positivity as opposed to daily clinical new cases. As such, this study compares the SARS-CoV-2 viral signal in wastewater to the percent positivity data acquired at the geographic region. The studied rural community (2,500-5,000 people) represents approximately 2% of the whole population within the health unit’s geographical boundaries for which epidemiological data are available. Although travel between communities in this geographic area is common, it is noted that presentation of the community SARS-CoV-2 viral signal against the epidemiological data at the geographic region level assumes that this town and its citizens behave similar to the residents of the larger region, which may not necessarily be a correct assumption. However, due to the limitations of available epidemiological information no other data were available to perform a comparison. As outlined by these results, it appears that samples harvested from the pumping station of the community could provide similar viral signal trends to the clinical data sets, making the pumping station a suitable sampling location for rural communities that are serviced by wastewater treatment lagoons. It is hypothesized that samples collected from the last pumping station of a smaller rural community will have similar usability for wastewater surveillance to samples collected from influent of a larger mechanical wastewater treatment plant.

When comparing the pumping station samples expressed as PMMoV-normalized viral genomic copies, viral genomic copies per gram of wastewater solids and viral genomic copies per L to available epidemiological data (clinical test percent positivity for the whole population) (Figure 4), a degree of visible agreement could be seen between the SARS-CoV-2 viral signal in wastewater and the reported percent positivity in the geographic region. However, this comparison may not be of significant relevance as the clinical data references a larger geographical zone than the sampled sewershed. It has been outlined in several studies that mass or volumetric normalization of SARS-CoV-2 viral signal alone may not truly capture sewershed dilution effects (Bivins et al., 2021; D’Aoust et al., 2021b; Wu et al., 2020). Furthermore, it is believed that while not observed or demonstrated with solid mass (copies/g) or volume (copies/L) normalized SARS-CoV-2 viral genomic copies, PMMoV-normalized viral genomic copies show a true change in community prevalence as the normalization with PMMoV helps account for the intrinsic quantity of fecal material in the wastewater (Graham et al., 2020; Kitamura et al., 2021; Wolfe et al., 2021; Wu et al., 2021). PMMoV normalization in WWS applications is believed to be particularly important in the context of surveillance of fecally shed pathogens as it allows to call true localized changes in prevalence which could otherwise be missed without normalizing for the quantity of fecal material in wastewater.

**Figure 3:**
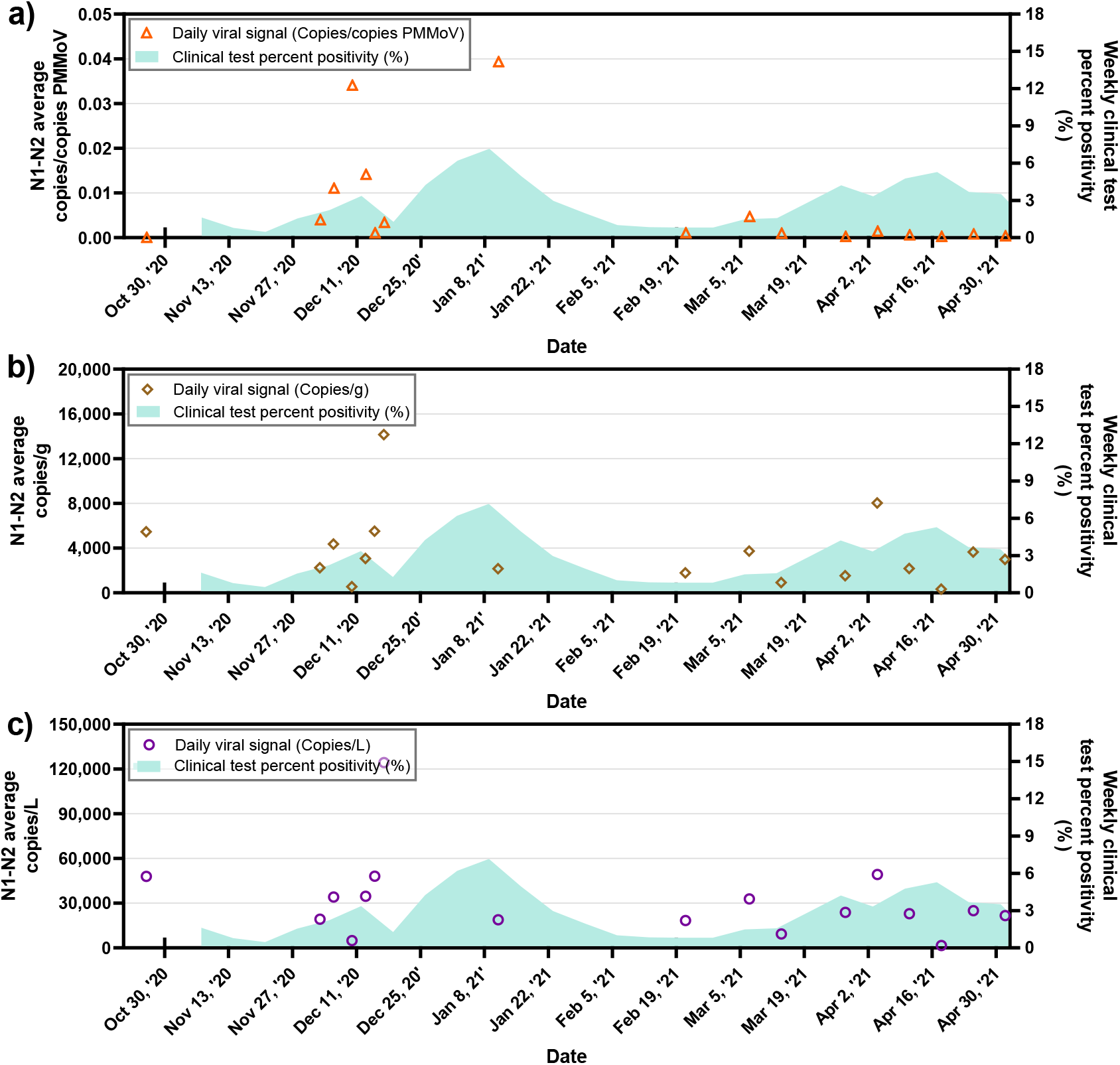
Average SARS-CoV-2 signal in the pumping station wastewater samples expressed as a) PMMoV normalized SARS-CoV-2 viral genomic copies, b) SARS-CoV-2 viral copies/g and c) SARS-CoV-2 viral genomic copies/L, along with the weekly COVID-19 test percent positivity.

## 4. Conclusions and recommendations

As many locations throughout the world have experienced new resurgence in COVID-19 cases due to the rapid spread of infectious variants of concern, it has become increasingly important for communities to implement effective COVID-19 wastewater monitoring programs that can rapidly detect and predict upcoming resurgences in COVID-19 cases. WWS programs may provide epidemiological information at a higher level of granularity to public health units than what is currently available in small, rural communities and low-income countries. Unfortunately, such programs are not frequently found in smaller, rural communities or low-income countries, and these communities often rely on low-cost, more passive treatment systems, compounding the gap of knowledge between the application of WWS to high-income, peri-urban and urban communities compared to low-income and rural communities. These smaller communities are particularly vulnerable to changes in funding or mandates as they may not have the resources to take over or start WWS initiatives on their own.

In this study, it was observed that in municipalities with wastewater lagoons, surveillance of changes in incidence of COVID-19 in the general population is possible via sampling from an upstream pumping station within the sewershed. Measurements of N1 and N2 SARS-CoV-2 gene regions and PMMoV in wastewater demonstrate consistent strong detection in samples collected at the upstream pumping station, but not from samples collected in wastewater treatment lagoons. Preliminary results show that samples collected from the wastewater treatment lagoon dosing polyaluminum sulphate for phosphorus removal demonstrates significant weaker detection due to potential preferential partitioning of SARS-CoV-2 viral particles to solids which rapidly settle upon entering the lagoon inlet structure. Furthermore, degradation of SARS-CoV-2 and PMMoV genetic material in the lagoon may be associated with the long retention time of the system. Finally, UV light exposure and degradation of the genetic material may also be a potentially significant mechanism of genetic material degradation. Even though SARS-CoV-2 genetic material is largely attached/associated to solids, the long retention time of the lagoon system may enable UV degradation of the solids partitioned material across a significant period of time. Additionally, it is observed that PMMoV-normalized viral signal may allow for detection of true increases in prevalence by normalizing measurements for the quantity of fecal material in wastewater. By observing PMMoV viral signal in wastewater, it may also be possible to distinguish SARS-CoV-2 N1 and N2 non-detects from false-negatives caused by sample degradation or severe inhibition. Finally, it is observed that wastewater-acquired SARS-CoV-2 viral signal from the upstream pumping station can be measured without difficulty and as such, could be used effectively in tandem with local epidemiological data to help detect and track new and existing COVID-19 outbreaks, even where localized increases in COVID-19 prevalence would not be visible with low granularity publicly available weekly regional epidemiological information.

## Data Availability

Data is fully available upon request.

## Acknowledgements

The authors wish to acknowledge the help and assistance of the University of Ottawa, the Ottawa Hospital, the Children’s Hospital of Eastern Ontario, the Children’s Hospital of Eastern Ontario’s Research Institute, Public Health Ontario and all their employees involved in the project. Their time, facilities, resources, and feedback are greatly appreciated. The authors also wish to specifically outline the assistance of Mr. Alain Castonguay.

